# Point prevalence of SARS-CoV-2 and infection fatality rate in Orleans and Jefferson Parish, Louisiana, May 9-15, 2020

**DOI:** 10.1101/2020.06.23.20138321

**Authors:** Amy K Feehan, Daniel Fort, Julia Garcia-Diaz, Eboni Price-Haywood, Cruz Velasco, Eric Sapp, Dawn Pevey, Leonardo Seoane

## Abstract

Using a novel recruitment method to reduce selection bias with paired molecular and antibody testing for SARS-CoV-2 infection, we determined point prevalence in a racially diverse municipality. Infections were highly variable by ZIP and differed by race. Overall census-weighted prevalence was 7.8% and the calculated infection fatality rate was 1.63%.

## Text

Seroprevalence studies around the world have estimated the spread of the novel severe acute respiratory syndrome coronavirus 2 (SARS-CoV-2) virus, but none have been performed in New Orleans, Louisiana, USA, an early epicenter of the outbreak. Additionally, COVID-19 has been widely reported to disproportionately affect Black patients. While the absolute number of deaths have been reported by race, we do not know the infection fatality rate (IFR) which requires knowing how many people are at risk (e.g. infected). This study was designed to estimate SARS-CoV-2 infections in Orleans and Jefferson Parishes (O/JP) and the COVID-19 related IFR by race.

The protocol was approved by the Ochsner IRB and designed to enroll and test up to 3,000 subjects at 10 sites throughout O/JP between May 9 and May 15, 2020. To recruit a representative sample for this high-throughput method, a novel two-step system developed by Public Democracy (Arlington, VA) considered more than 50 characteristics, including social determinants of health and US Census population data, to establish a pool of potential participants reflective of the demographics of the Parishes, from which a randomized subset of 150,000 was selected.

Over 25,000 volunteers were recruited from this subset through dynamic, cross-device digital ads. This volunteer pool was stratified by the same attributes and then randomly issued a text message inviting subjects to private testing locations. Invitations were adjusted daily based on response rates to ensure we achieved our *a priori* goals for a representative sample. Volunteers checked in with a QR code or phone number to discourage walk-ups. Ochsner Health did not turn uninvited people away but excluded these from analysis if they did not fit criteria. Family members of participants (234), or people who lived in ineligible ZIP codes (34) were excluded. Six people withdrew consent. Digital ads, consent forms, and surveys were created in English, Spanish, and Vietnamese. Participants were offered free taxi service to and from the test sites. Verbal consent was electronically documented, and subjects were asked a short list of questions followed by a blood draw and nasopharyngeal (NP) swab.

US Food and Drug Administration-Emergency Use Authorization approved tests were used. Real-time reverse transcriptase polymerase chain reaction (PCR) tests of NP swabs were performed on the Abbott m2000 RealTime system. Qualitative Immunoglobulin G (IgG) blood tests were performed on the ARCHITECT i2000SR. The IgG test meets criteria described by the CDC to yield high positive predictive value, which was validated by Ochsner Health laboratory and by others. (*1,2*) Study participants with either or both positive tests were assessed as having been infected with SARS-CoV-2. Census-weighted values were calculated for prevalence and seroprevalence. The positive-testing population included early-stage infections (PCR+ only) as well as people recovering (PCR+ and IgG+) and recovered (IgG+ only). Early-stage infections were excluded from IFR estimation as the outcomes of their infections would not yet be registered as official deaths. Therefore, weighted seroprevalence (anyone with an IgG+ result) was used to calculate “presumed recovered.” IFR was calculated by dividing cumulative deaths by race, reported by the Louisiana Department of Health, by “presumed recovered” individuals.

Among the 2,640 analyzed, the sample was 63.5% female, 61.8% white, average age of 50.6 years, and average household size of 2.55 people. Among the 183 who tested positive, 49% were Black. The raw prevalence of SARS-CoV-2 in the sample population is 6.9% (7.8%, census-weighted) with 2% positive for active viral shedding (PCR+, with or without IgG). By race, prevalence was highest (10.3%) in Black subjects followed by multiracial (9.4%), Asian (6.4%), and white (5.9%). Hispanic prevalence was 7.5%. 2018 population estimates are indicated in Table 1 and multiplied by weighted seroprevalence (percent IgG+) to generate the number of “presumed recovered.” Reported deaths are divided by “presumed recovered” to calculate the IFR, which was 1.63% overall. The IFR was statistically similar for white (1.58%), Black (1.72%), and multiracial (1.40%), but Asian IFR was significantly lower (0.61%). Hispanic deaths are not currently reported for O/JP.

**Table 1.** Prevalence and IFR in Orleans and Jefferson Parish after 7 weeks of an active stay-at-home order.

|  | Positive<br>n (total) | Number of<br>O/JP<br>Residents <sup>a</sup> | Raw Prevalence<br>% (CI) | Weighted<br>Prevalence <sup>b</sup><br>% (CI) | Weighted<br>Seroprevalence<br>% (CI) | Presumed<br>recovered <sup>c</sup> | Deaths as<br>of May<br>16, 2020 <sup>a</sup> | IFR<sup>d</sup><br>% (CI) |
| --- | --- | --- | --- | --- | --- | --- | --- | --- |
| Total | 183 (2640) | 825,057 | 6.9 (6.0,8.0) | 7.8 (7.76,7.88) | 6.86 (6.8,6.91) | 56,578 | 925 | 1.63 (1.53,1.74) |
| White | 79 (1607) | 419,800 | 4.9 (3.9,6.1) | 5.9 (5.78,5.93) | 4.5 (4.44,4.58) | 18,975 | 299 | 1.58 (1.41,1.77) |
| Black | 90 (828) | 356,925 | 10.9 (8.8,13.2) | 10.3 (10.19,10.39) | 9.8 (9.7,9.9) | 34,973 | 600 | 1.72 (1.58,1.86) |
| Asian | 9 (130) | 29,740 | 6.9 (3.2,12.7) | 6.4 (6.13,6.69) | 5.5 (5.2,5.7) | 1,629 | 10 | 0.61 (0.33,1.14)<sup>e</sup> |
| Native American | 0 (14) | 4,088 | 0 | 0 | 0 | - | 0 | - |
| Pacific Islander | 0 (3) | 495 | 0 | 0 | 0 | - | 2 | - |
| Multiracial /other | 5 (58) | 14,009 | 8.6 (2.9,19.0) | 9.4 (8.96,9.94) | 7.1 (6.7,7.6) | 1,001 | 14 | 1.40 (0.83,2.36) |
| Hispanic <sup>f</sup> | 18 (293) | 86,289 | 6.1 (3.7,9.5) | 7.5 (7.30, 7.65) | 5.3 (5.16,5.46) | 4,582 | unknown | - |

<sup>a</sup> 2018 population estimates and deaths by race reported by ldh.la.gov
<sup>b</sup> Census-weighted prevalence of PCR+ and/or IgG+ tests calculated to match 2018 demographics by Parish and then combined.
<sup>c</sup> Number of residents multiplied by weighted seroprevalence (IgG+ tests).
<sup>d</sup> Infection Fatality Rate (IFR) equals the number of deaths over presumed infections.
<sup>e</sup> Significantly lower than white (p=0.0034), Black (p=0.0013) and multiracial (p=0.0467)
<sup>f</sup> Hispanic ethnicity is a separate analysis and numbers were not subtracted from race. Hispanic deaths were not reported by the state as of May 16, 2020.

The prevalence of viral shedding (PCR+) and overall SARS-CoV-2 exposure (IgG+) are listed and mapped by ZIP codes across O/JP in Figure 1. Prevalence was highly variable across the map and in some areas >20%.

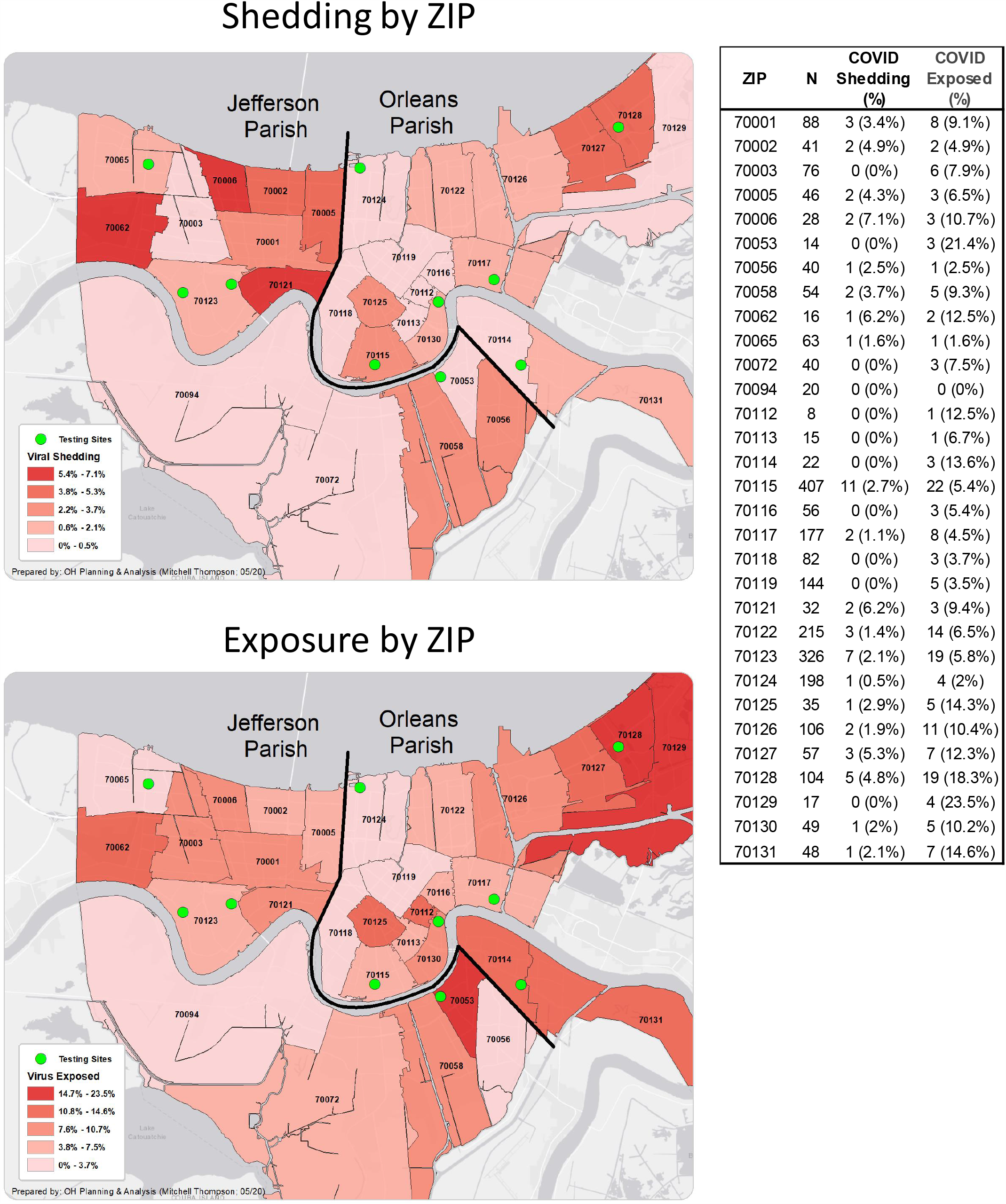

Prevalence studies help to understand infection spread, especially when testing resources are limited. This representative, high minority enrollment study found an overall prevalence of PCR+ and/or IgG+ tests to be 7.8%. Hispanics and Asians had higher prevalence compared to whites, and this study confirms a recent report of over-representation of Black individuals with COVID-19 infection in the New Orleans area. (*4*) The overall IFR was 1.63%, which agrees with a recent estimate of 1.3% IFR (0.6%, 2.1%) for the US (*3*). The similar IFR among most racial groups indicates that viral spread at least partially explains the increased number of deaths among minorities.

## Data Availability

De-identified data is available upon reasonable request to the corresponding author.

## Acknowledgments

The authors would like to especially thank the labs at the Ochsner Medical Center Jefferson Highway Campus for testing and keeping track of research samples; Dan Nichols, marketing technologist at Public Democracy; George Hutter and ReNOLA for their financial support; Sarah Roberts and Gina Mmahat for clinical site management; Samantha Bright, Lyndsey Buckner-Baiamonte, and Ansley Hammons for research site management; Emily Arata for liaising with public leaders; and countless research coordinators, clinical staff, marketing personnel, medical students, and Epic and IT staff for making site testing possible. The Ochsner Health Market Planning and Analysis team designed the maps in Figure 1. The authors thank Kathleen McFadden for her thorough editing and Dr. Mark Roberts for his review. We would also like to acknowledge the New Orleans Mayor’s Office, City of New Orleans Office of Public Health, and the New Orleans City Council, especially council members Helena Moreno and Jason Williams for filming a public service announcement to help recruit participants. We also thank Jefferson Parish President Cynthia Lee Sheng and Parish Council for their support.

## Notes

### Competing Interest Statement

A charitable donation was made by ReNOLA, a non-profit, to cover the cost of recruitment by Public Democracy, owned and operated by author Eric Sapp. No other authors have competing interests.

### Funding Statement

ReNOLA funded the recruitment effort by Public Democracy but was not involved in the study design, execution, or analysis. Ochsner Health funded the remainder of the study and all authors, except for Eric Sapp, are employed by Ochsner Health. Therefore, Ochsner Health was involved in the design, execution, analysis, and writing of this paper. The authors have not been paid to write this article beyond regular employment contracts.

### Author Declarations

The Ochsner Health IRB approved this study and can be contacted at or by phone at 504-842-3535.

